# GWAS identifies genetic loci for antibody response to SARS-CoV-2 vaccines in patients with systemic autoimmune diseases and healthy individuals

**DOI:** 10.1101/2025.05.16.25327770

**Authors:** Kwangwoo Kim, Dillon Claybaugh, Eduardo Patino-Martinez, Yenealem Temesgen-Oyelakin, Elaine Poncio, Jun Chu, Michael Davis, Alice Fike, Yanira Ruiz-Perdomo, Julie Onyechi, Margaret Beach, Lilian Howard, Eileen Pelayo, Nancy Spencer, Martha Sully, Rita Volochayev, Sophie Kelly, Sarah Porche, Laura B. Lewandowski, Luis M. Franco, Zerai Manna, Sarthak Gupta, Amy Hutchinson, Lisa Mirabello, Vibha Vij, Kaitlin A. Quinn, Peter C. Grayson, Adam Schiffenbauer, Lisa G. Rider, Iago Pinal-Fernandez, Andrew L. Mammen, Heather R. Kalish, Meryl A. Waldman, Blake Warner, Sarfaraz Hasni, Stephen J. Chanock, Mariana J. Kaplan

## Abstract

The efficacy of nucleic acid-based vaccines against SARS-CoV-2 varies across individuals, partly due to genetic factors influencing neutralizing antibody production. In patients with systemic autoimmune diseases (SADs), this response may be further altered by immune dysregulation. We conducted a genome-wide association study (GWAS) to identify genetic variants associated with post-vaccination anti-SARS-CoV-2 IgG antibody levels and to assess whether these associations differ between SAD patients and healthy individuals. The study included 165 participants (138 with SADs, 27 healthy controls), all of whom received nucleic-acid based vaccines. Antibody levels targeting the spike protein receptor-binding domain (RBD) and nucleocapsid were measured between one and twelve months after vaccination. GWAS results were meta-analyzed with data from a previously published GWAS of 1,076 healthy individuals. We identified a novel association near *RACGAP1* (rs706785; βmeta=–0.30, Pmeta=3.85×10−□) and replicated a known association at HLA-DRB1 position 71 (βmeta=–0.23, Pmeta=1.94×10^−11^). No significant interactions were observed between genotype and disease status. This study highlights both MHC and non-MHC genetic contributions to SARS-CoV-2 vaccine responses and suggests these effects are consistent across SAD patients and healthy individuals, supporting standard vaccination strategies for individuals with systemic autoimmune conditions.

## INTRODUCTION

The global impact of the severe acute respiratory syndrome coronavirus 2 (SARS-CoV-2) pandemic has underscored the critical importance of effective vaccines in controlling the spread of the virus and mitigating severe disease outcomes. Vaccines developed using mRNA and adenoviral vector technologies have shown high efficacy in preventing severe COVID-19.^1^ A key factor in this efficacy is the level of neutralizing antibodies that target the viral spike protein, particularly its receptor-binding domain (RBD).^2,3^ However, there is considerable variability in the levels of neutralizing antibodies among individuals after vaccination, which may be influenced by a range of factors, including genetic predisposition. Indeed, multiple genome-wide association studies (GWAS) have identified genetic association signals within the major histocompatibility complex (MHC) region, especially with the most significant association at HLA-DRB1 amino acid position 71.^4,5^

Variability in antibody responses could be even more pronounced in patients with systemic autoimmune disorders (SAD), who may exhibit altered antibody production due to preexistent immune dysregulation. Genetic variants affecting antibody levels in these patients could have distinct effect sizes compared to those in healthy individuals. Moreover, if variants associated with SAD risk influence antibody levels, it could suggest a need for tailored vaccine protocols to optimize efficacy for SAD patients.

This study aimed to identify genetic variants associated with antibody levels after SARS-CoV-2 nucleic acid vaccination through a GWAS involving SAD patients and healthy controls. Additionally, we investigated whether SAD diagnosis modifies the effect of these genetic variants by examining disease-SNP interactions.

## PATIENTS AND METHODS

### Subjects and vaccination

The analysis-ready sample comprised 165 participants, including patients with systemic lupus erythematosus (SLE, *n*=61), rheumatoid arthritis (RA, *n*=29), immune-mediated kidney disease (KD, *n*=11), Sjögren’s syndrome (SS, *n*=19), idiopathic inflammatory myopathies (IIM, *n*=9), and ANCA-vasculitis (*n*=9), as well as 27 healthy controls. All SAD patients fulfilled established criteria for their specific systemic autoimmune diseases. The IgG antibody levels against the RBD and nucleocapsid of SARS-CoV-2 were measured in heat-inactivated serum samples using a previously optimized enzyme-linked immunosorbent assay (ELISA)-based approach^6^, between 28 and 365 days after the last vaccination. All participants received COVID-19 mRNA vaccines, specifically produced by Pfizer–BioNTech (BNT1622b2) or Moderna (Spikevax), with at least two doses of the same vaccine. Regarding vaccination doses, 125 participants received two doses, 36 received three doses, and 4 received four doses before antibody measurements. Patients were recruited at the NIH Clinical Center and provided informed written consent to participate. This study was approved by the NIH Institutional Review Board (IRB-000207 COVID-SAD). This study adhered to all relevant ethical regulations, including compliance with the Declaration of Helsinki

### Genetic data

DNA was purified from peripheral blood. Genotype data were generated at the Cancer Genomics Research Laboratory of DCEG-NCI as part of the COVNET consortium (https://dceg.cancer.gov/research/how-we-study/genomic-studies/covnet) on an Infinium Global Screening Array (GSAMD-24v2-0), which includes 712,189 markers. To ensure high-quality genetic data, several criteria were applied for variant exclusion. Variants with low minor allele counts (MAC<4; *n*=177,431) and low call rates (<97%; *n*=21,035) were excluded. Indels and G/C or T/A SNPs (4,035 variants) were removed, as well as duplicates (1,482 variants). After inferring ancestral origin of each genetic variant for each individual (described below), variants exhibiting Hardy-Weinberg disequilibrium (*n*=711 variants; *P*_HWE_<0.001 in any ancestry or *P*_HWE_<0.01 in more than two ancestries) were also excluded. Sample quality checks confirmed that all subjects demonstrated high genotyping call rates, no relatedness (up to the third degree), and consistency between self-reported and genetic sex.

Whole-genome imputation was performed using Beagle5,^7^ with reference phases derived from the 1,000 Genomes Project (1KGP) phase 3. Post-imputation quality control retained imputed variants with minor allele frequencies (MAFs) ≥ 0.005 and imputation quality scores (R^2^) ≥ 0.7, yielding a total of 13,566,613 variants across 165 samples. HLA imputation was conducted using Eagle v2.4 and Minimac4 with a high-resolution multi-ethnic HLA imputation panel to impute HLA classical alleles and amino acid residues.^8^

### Local ancestry inference

We performed ancestry-specific segment analysis using RFMix v2 (https://github.com/slowkoni/rfmix) to assign local ancestry segments along fully phased imputed data for each study subject, based on ancestry-informed reference haplotypes from five target ancestries: European, African, Latino/Admixed American, Central/South Asian, and East Asian.

For the ancestry reference haplotypes, we curated a set of 3,593 individuals from the gnomAD HGDP/1KGP callset (gnomAD v3.1.2; https://gnomad.broadinstitute.org/downloads), filtering to include only individuals with an ancestry inference probability of 1 for one of the target ancestries. Ancestry inference probabilities for the gnomAD HGDP/1KGP callset were previously estimated (https://gnomad.broadinstitute.org/news/2021-10-gnomad-v3-1-2-minor-release/) and are available through the gnomAD project. For HLA imputation data, we constructed a separate ancestry reference dataset using the same gnomAD HGDP/1KGP samples, applying HLA imputation methods as described earlier.

This approach enabled high-resolution, ancestry-specific calls across the genome and within the MHC region, allowing us to calculate ancestry-specific haplotype dosage at each variant and the ancestry-specific dosage of its effect allele in everyone.

### Genetic association analysis

The genetic association analysis was conducted using a multivariate linear regression framework, as described in the Tractor method,^9^ to deconvolute ancestry-specific effect sizes for each variant. The association model applied for each variant is as follows:

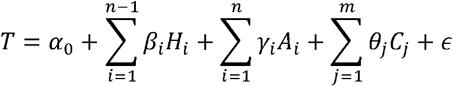

where *T* represents the normalized antibody titer for anti-RBD IgG; *H*_*i*_ is the dosage of haplotypes from the *i*-th ancestry; *A*_*i*_ is the dosage of effect alleles from the *i*-th ancestry; and *C*_*j*_ denotes the *j*-th covariate, which includes the top five genetic principal components (PCs), sex, age, SAD diagnosis, log-transformed days between last vaccination and antibody measurement, vaccine manufacturer, number of vaccine doses, and normalized anti-nucleocapsid IgG titer. In this model, *α*_0_ is the intercept term, *β*_*i*_ is the ancestry-specific effect estimate of haplotype dosages, *γ*_*i*_ is the ancestry-specific effect estimate for a given genetic variant, *θ*_*j*_ represents the effect estimate for the *j*-th covariate, and *ϵ* is the error term. This model includes ancestry-specific haplotype dosages from n-1 ancestries to avoid collinearity. Antibody titers were transformed using a rank-based inverse normal transformation, which was applied to both anti-RBD and anti-nucleocapsid titers. This transformation was essential for conducting meta-analysis with previously published datasets.

The deconvoluted ancestry-specific effect sizes (*γ*) for each variant were subsequently aggregated using inverse-variance-weighted meta-analysis. Effect sizes from ancestries with a haplotype dosage below 5%, an MAF less than 5%, or an MAC below 3 were excluded from the meta-analysis. The aggregated effect sizes were then meta-analyzed with results from a previous GWAS conducted on 1,076 individuals in a UK vaccine efficacy trial.^5^ In this external dataset, IgG RBD levels, normalized using a rank-based inverse normal transformation, were assessed 28 days after initial vaccination with the AstraZeneca AZD1222 adenoviral vector vaccine encoding the spike protein.

For variants reaching genome-wide significance for antibody titers, we investigated potential interactions between genetic variants and disease diagnosis. We compared model fits between the regression model described above and an expanded model that additionally included interaction terms, as follows:

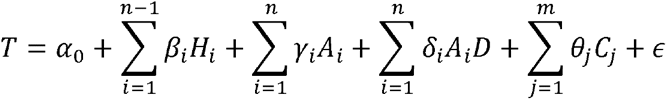

where *δ*_*i*_ represents the ancestry-specific effect estimate for the interaction between a genetic variant and SAD diagnosis in the *i*-th ancestry. Disease status *D* was encoded as 1 for SAD patients and 0 for healthy controls.

## RESULTS

We analyzed over 13 million autosomal variants from whole-genome imputation along with HLA variants, including classical alleles and amino acid residues, in a cohort of 165 individuals (138 SAD patients and 27 healthy controls). Our study cohort included participants from diverse ancestries, and some individuals exhibited mixed genetic ancestry. To adjust for ancestral background in our association analysis, we conducted variant-level local ancestry inference for everyone using RFMix v2, with the gnomAD HGDP/1KGP callset as the reference dataset (Figure 1a). Ancestry proportions inferred from this process were associated with the top three genetic PCs, which explained substantial portions of the genetic variance in the study cohort (Figures 1b and 1c).

**Figure 1.**
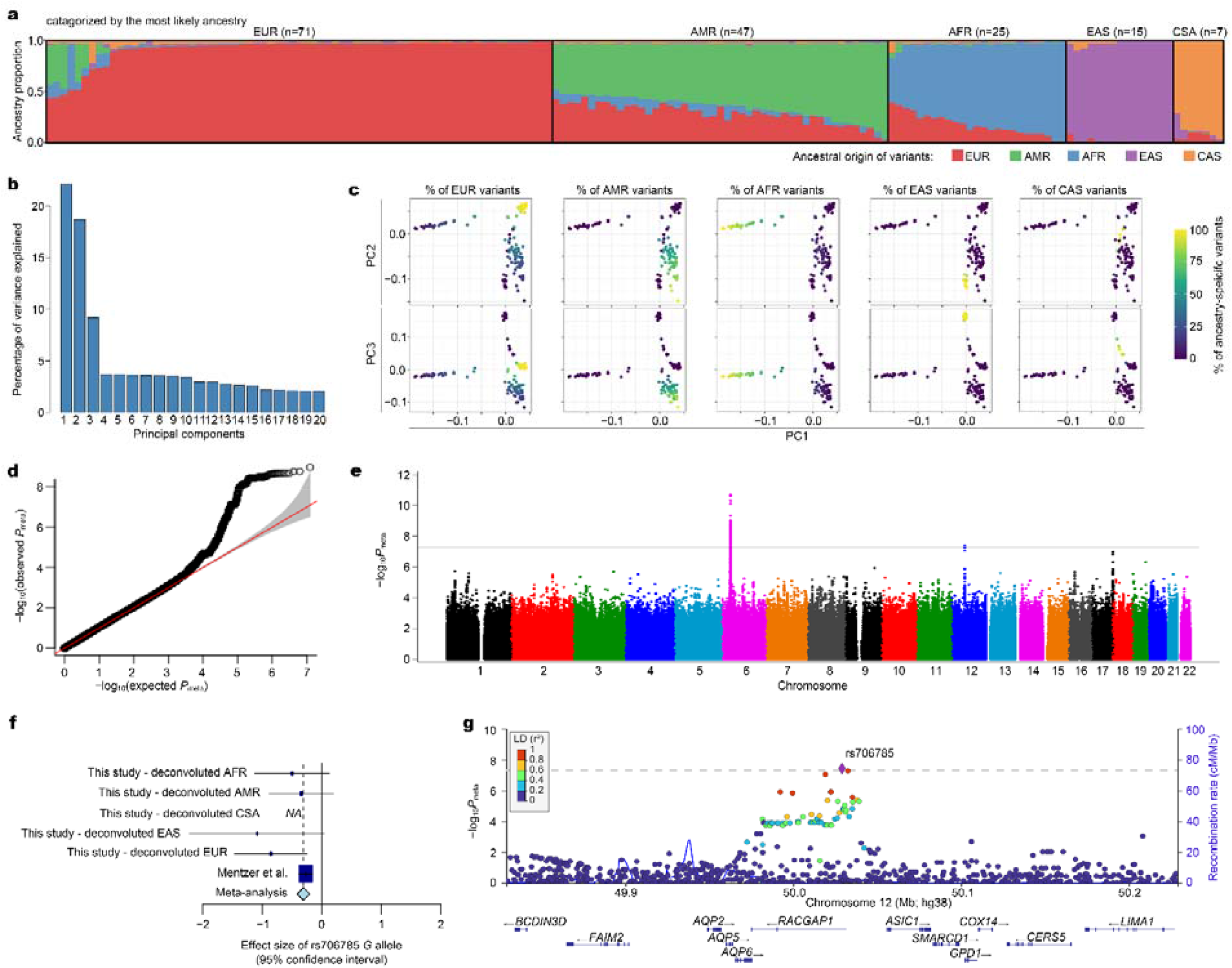
**(a)** Ancestral composition of study participants (*n*=165) across five major population groups: European (EUR), African (AFR), Latino/Admixed American (AMR), Central/South Asian (CSA), and East Asian (EAS). Vertical bars represent genetic ancestry proportions estimated using local ancestry inference. **(b)** Relative variance explained by the top 20 PCs in the study cohort. **(c)** Scatter plots of PC1, PC2, and PC3, with data points color-coded by individual’s ancestry-specific proportions, highlighting the clustering of participants by genetic ancestry. **(d)** Quantile-quantile plot of *P* values from the meta-analysis combining this study and the data from Mentzer *et al*. The red diagonal line represents perfect agreement between observed and expected distributions, with the 95% confidence interval shaded in gray. **(e)** Manhattan plot of –log_10_*P*_meta_ (y-axis) for each variant according to chromosomal position (x-axis). The genome-wide significance threshold is denoted by a horizontal gray line. **(f)** Forest plot for rs706785, depicting the effect sizes (dots) of the *G* allele with 95% confidence intervals across deconvoluted ancestries, the prior GWAS, and the meta-analysis. **(g)** Regional association plot for the *RACGAP1* locus, displaying –log_10_*P*_meta_ by chromosomal position. Variants are color-coded based on their LD with the most significant variant (rs706785), based on the 1KGP EUR reference panel.

Genome-wide association analysis was performed to estimate ancestry-specific effect sizes for each variant. We then meta-analyzed these deconvoluted effect sizes to obtain global effect estimates, adjusting for various covariates described in our methods section, including genetic PCs (PC1-PC5), sex, SAD or control diagnosis, vaccine dose, vaccine manufacturer, and time since the last vaccination (in log-transformed days), as well as anti-nucleocapsid IgG antibody levels.

Among these covariates, time since vaccination and anti-nucleocapsid IgG levels showed significant associations with antibody titers (Table 1). Notably, the log-transformed days since the last vaccination were inversely associated with antibody levels, suggesting a decline in titers over time. In contrast, anti-nucleocapsid IgG antibody levels, used as an additional control for SARS-CoV-2 exposure, were significantly associated with higher levels of anti-RBD IgG antibodies. Although the comparison did not reach significance after accounting for multiple testing, individuals who received the Moderna vaccine tended to have higher antibody titers compared to those who received the Pfizer vaccine, which is consistent with expectations due to the higher mRNA dose in the Moderna vaccine.

**Table 1.**
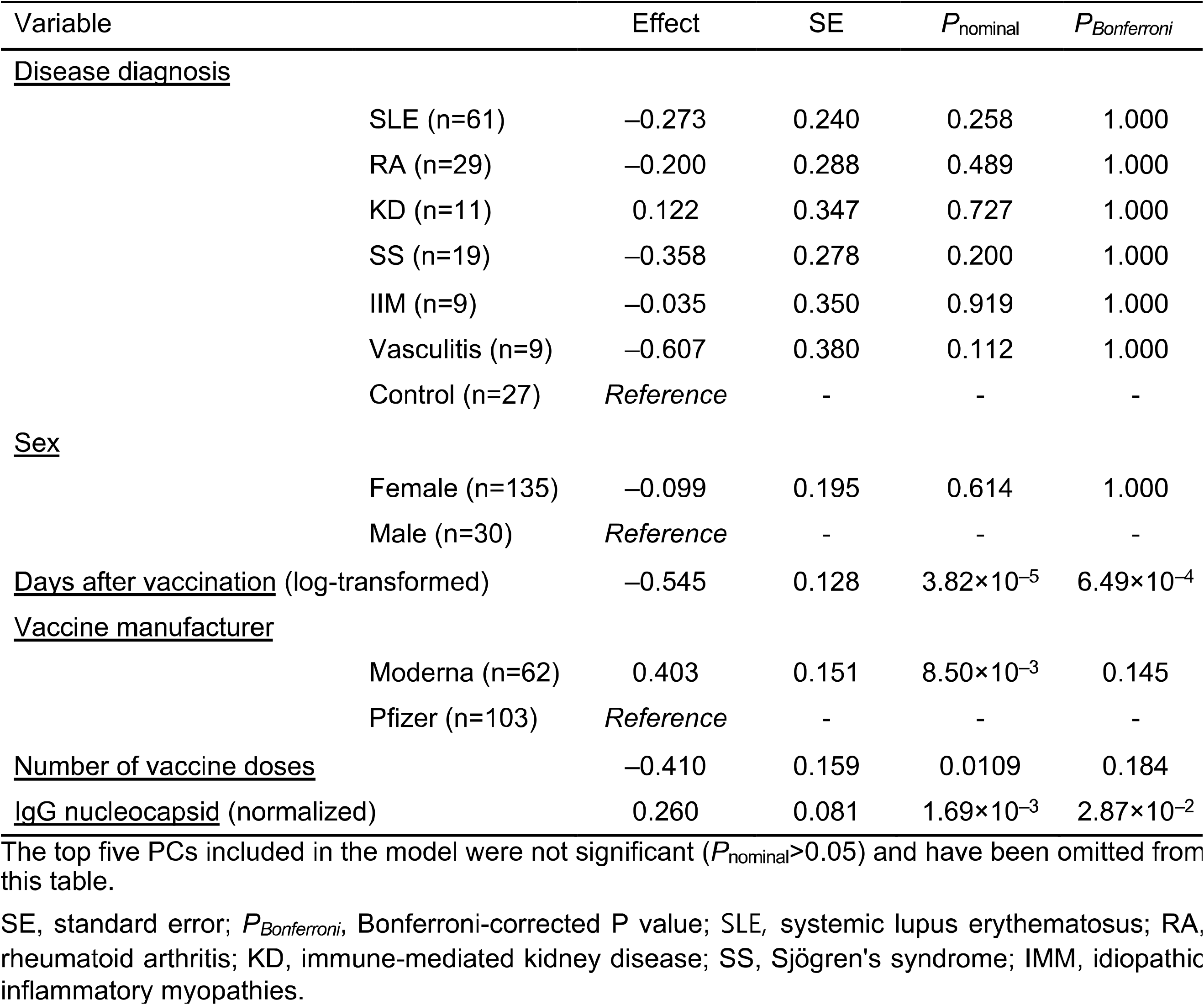
Multivariate linear regression for the normalized level of anti-SARS-CoV-2 RBD lgG antibody.

We conducted GWAS using variant-level local ancestry information to estimate ancestry-specific effect sizes of genetic variants in multivariate linear regression and to estimate combined effect sizes through inverse-variance-weighted meta-analysis (see Methods). The genomic inflation factor (λ) from our genome-wide association results was 1.05, indicating minimal systemic bias and no evidence of significant underlying population substructure. We found no variants surpassing the genome-wide significance threshold (*P*=5×10^−8^), likely due to the limited sample size of the study.

To improve statistical power and result robustness, we conducted a meta-analysis by integrating our data with results from a recent GWAS^5^ that employed the same normalized antibody measurement method, enabling comparability of genetic effect sizes across studies. The genomic inflation factor (λ) was 1.06 (Figure 1d). In this meta-analysis, we replicated the previously reported association of HLA-DRB1 amino acid position 71 with antibody titers (*P*_meta_=1.94×10^−11^; Table 2; Figure 1e), achieving a slightly stronger significance than the original study. The presence of arginine or serine at this position was associated with lower antibody titers (β_meta_=– 0.23), with consistent findings in both our study (β=–0.25, *P*=2.63×10^−2^) and the previous GWAS (β=–0.23, *P*=2.38×10^−10^). Notably, position 71 is a critical site that shapes the epitope-binding surface of HLA-DR molecules and determines the “shared epitope” associated with RA.^10^ Arginine at this position is not exclusively present in shared epitope alleles nor associated with RA risk.^11^ Serine at position 71 is very rare in multiple populations.

**Table 2.**
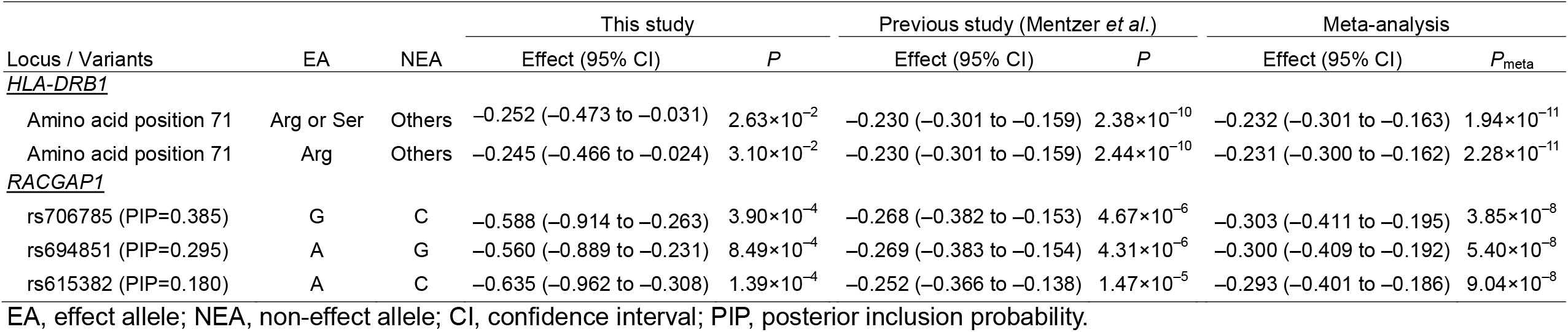
Association of HLA-DRB1 and RACGAP1 variants with post-vaccination anti-SARS-CoV-2 IgG antibody levels.

| Locus / Variants | EA | NEA | This study |  | Previous study (Mentzer *et al.*) |  | Meta-analysis |  |
| --- | --- | --- | --- | --- | --- | --- | --- | --- |
|  |  |  | Effect (95% CI) | *P* | Effect (95% CI) | *P* | Effect (95% CI) | *P*<sub>meta</sub> |
| <u>HLA-DRB1</u> |  |  |  |  |  |  |  |  |
| Amino acid position 71 | Arg or Ser | Others | −0.252 (−0.473 to −0.031) | 2.63×10<sup>−2</sup> | −0.230 (−0.301 to −0.159) | 2.38×10<sup>−10</sup> | −0.232 (−0.301 to −0.163) | 1.94×10<sup>−11</sup> |
| Amino acid position 71 | Arg | Others | −0.245 (−0.466 to −0.024) | 3.10×10<sup>−2</sup> | −0.230 (−0.301 to −0.159) | 2.44×10<sup>−10</sup> | −0.231 (−0.300 to −0.162) | 2.28×10<sup>−11</sup> |
| <u>RACGAP1</u> |  |  |  |  |  |  |  |  |
| rs706785 (PIP=0.385) | G | C | −0.588 (−0.914 to −0.263) | 3.90×10<sup>−4</sup> | −0.268 (−0.382 to −0.153) | 4.67×10<sup>−6</sup> | −0.303 (−0.411 to −0.195) | 3.85×10<sup>−8</sup> |
| rs694851 (PIP=0.295) | A | G | −0.560 (−0.889 to −0.231) | 8.49×10<sup>−4</sup> | −0.269 (−0.383 to −0.154) | 4.31×10<sup>−6</sup> | −0.300 (−0.409 to −0.192) | 5.40×10<sup>−8</sup> |
| rs615382 (PIP=0.180) | A | C | −0.635 (−0.962 to −0.308) | 1.39×10<sup>−4</sup> | −0.252 (−0.366 to −0.138) | 1.47×10<sup>−5</sup> | −0.293 (−0.401 to −0.186) | 9.04×10<sup>−8</sup> |

EA, effect allele; NEA, non-effect allele; CI, confidence interval; PIP, posterior inclusion probability.

Additionally, in a meta-analysis, we identified a novel association locus near the *RACGAP1* gene (Figures 1e, 1f, and 1g). The most significant association was detected at rs706785, located 3.5 kb upstream of *RACGAP1* (β_meta_=–0.30; *P*_meta_=1.94×10^−11^; Table 2; Figure 1g). Consistent effect sizes were observed across both our study (β=–0.59, *P*=3.90×10^−4^) and the prior GWAS (β=–0.27, *P*=4.67×10^−6^). To further investigate the locus, we conducted a fine-mapping analysis and identified six variants within the 90% credible set for this locus,^12^ based on approximate Bayesian factors with a prior variance of 0.04 in allelic effects. Among these, three variants located in the upstream and intronic regions of *RACGAP1* accounted for 86% of the posterior probability (Table 2). Notably, these three variants have been reported as expression quantitative trait loci (eQTLs) for multiple genes, including *RACGAP1, CERS5, LIMA1*, and *COX14*, across various tissues (Supplementary Table 1). This observation is also supported by chromatin interaction data, which suggests potential regulatory interactions within this locus (Supplementary Figure 1).For instance, the *G* allele of rs706785, associated with lower antibody titers, is also linked to decreased *RACGAP1* expression levels.

Although genetic variants of *RACGAP1* and an alanine or serine residue at HLA-DRB1 amino acid position 71 have not been associated with SAD, we investigated potential interactions of these variants with disease diagnosis to examine whether the observed GWAS effect sizes are distinct in patients with SAD compared to healthy individuals. Logistic regression analysis, incorporating interaction terms between disease diagnosis and ancestry-specific allelic dosages, did not reveal any significant interaction effects associated with antibody levels (*P*>0.05).

## DISCUSSION

In this study, we investigated the genetic underpinnings of antibody responses to nucleic acid vaccines against SARS-CoV-2 by performing a GWAS and a subsequent meta-analysis with a recent study that utilized the same antibody measurement method. We confirmed the previously reported association between HLA-DRB1 amino acid position 71 and anti-RBD IgG titers. Furthermore, we identified a novel signal near the *RACGAP1* gene, which includes eQTLs for *RACGAP1* and nearby genes, underscoring the potential role of this locus in vaccine-induced immunity.

By leveraging genetic data from participants of diverse ancestries, our analysis effectively accounted for ancestral differences and minimized bias through robust methodologies, including ancestry-specific effect size estimation and Bayesian fine-mapping. These approaches enhanced our ability to identify genetic variants associated with antibody responses, despite the relatively small sample size. The integration of our findings with data from a prior GWAS significantly increased statistical power, enabling cross-study validation and the discovery of a novel locus associated with antibody levels. This combined analysis not only reinforced the evidence for previously implicated genetic variants but also identified variants around *RACGAP1* as a potential contributor to the variability in vaccine-induced immune responses. Our results should be pursued in subsequent studies designed to identify and characterize further genetic variants that contribute to antibody responses to SARS-CoV-2 in individuals diagnosed with specific SADs, as well as interactions with immunosuppressive medications. Importantly, the inclusion of covariates such as vaccine manufacturer, number of doses, and time since vaccination allowed us to adjust for non-genetic factors that influence antibody titers, providing a clearer view of the genetic effects on SARS-CoV-2 vaccine responses.

We also investigated potential interactions between genetic variants and disease diagnosis. Our results did not reveal significant interactions, suggesting that the genetic variants associated with antibody responses operate similarly in both SAD patients and healthy individuals.

We identified several candidate genes regulated by credible set variants around *RACGAP1*, although direct evidence linking them to vaccine efficacy or antibody levels is currently lacking. Among these, *RACGAP1*, a member of the Rho GTPase-activating protein family, is primarily known for critical roles in cytokinesis, cell migration, and invasion, through its regulation of cytoskeletal dynamics and signaling pathways.^13,14^ While *RACGAP1* has been extensively studied in cancer biology— where its overexpression is associated with poor prognosis across various malignancies—its role in immune responses remains poorly understood. Recent findings correlating *RACGAP1* expression in various cancer cells with immune cell infiltration suggest a potential role in modulating immune cell dynamics and responsiveness.^15^ In the context of vaccine-induced immunity, which requires immune cell infiltration to trigger an effective immune response at the site of injection or in proximal lymph nodes, the observed association of *RACGAP1* expression-reducing alleles with lower antibody titers suggests that these alleles may weaken immune cell activation or recruitment, potentially dampening vaccine efficacy. Further functional studies and independent GWASs are needed to validate our findings and to clarify potential mechanisms and their implications for immune responses.

In conclusion, this study provides new insights into the genetic basis of vaccine-induced antibody responses, marking the discovery of the first non-MHC genetic association with SARS CoV-2 vaccine-induced immunity. Notably, the absence of detectable interactions between the identified genetic variants and disease diagnosis suggests that, at least for the loci investigated, the heritable genetic effects on vaccine-induced immune responses in SAD patients may be comparable to those observed in the general population.

## ACKNOWLEDGMENTS

The authors sincerely thank the patients who generously participated in this study. We are also deeply grateful to Dr. Alexander J. Mentzer from the Wellcome Centre for Human Genetics, Nuffield Department of Medicine, University of Oxford, Oxford, UK, for providing the GWAS summary statistics from his recent study. This work was supported by the NIH (Intramural Targeted anti-COVID Award from NIAID; the Intramural Research Program at NIAMS (ZIA AR-041199), NIDDK and NIEHS and with Federal funds from the National Cancer Institute, National Institutes of Health, under Contract No. 75N91019D00024. The content of this publication does not necessarily reflect the views or policies of the Department of Health and Human Services, nor does mention of trade names, commercial products, or organizations imply endorsement by the U.S. Government.).

## CONFLICT OF INTEREST

The authors report no conflicts of interest associated to this research.

## AUTHOR CONTRIBUTIONS

All authors contributed to at least one of the following manuscript preparation roles: conceptualization AND/OR methodology, software, investigation, formal analysis, data curation, visualization, and validation AND drafting or reviewing/editing the final draft. As corresponding author, Dr. Kaplan confirms that all authors have provided the final approval of the version to be published, and takes responsibility for the affirmations regarding article submission (eg, not under consideration by another journal), the integrity of the data presented, and the statements regarding compliance with institutional review board/Helsinki Declaration requirements.

## DATA AVAILABILITY

The genetic dataset generated and/or analyzed during the current study are not publicly available to protect privacy but are available from the corresponding author on reasonable request.

## Notes

### Competing Interest Statement

The authors have declared no competing interest.

### Author Declarations

This study was approved by the NIH IRB

