## Supplemental Table and Figures for "GWAS identifies genetic loci for antibody response to SARS-CoV-2 vaccines in patients with systemic autoimmune diseases and healthy individuals"

**Supplementary Material**

**Supplementary Table 1. Reported eQTL effects of credible set variants rs706785, rs694851, and rs615382**

| **rsID** | **DB*** | **Tissue*** | **Symbol** | **Effect allele** | **Stats** | **P** | **FDR-corrected P** |
| --- | --- | --- | --- | --- | --- | --- | --- |
| rs706785 | eQTLGen | eQTLGen_cis_eQTLs | *CERS5* | C | -15.10 | 1.7E-51 | 0 |
| rs706785 | eQTLGen | eQTLGen_cis_eQTLs | *RACGAP1* | C | 11.72 | 1.0E-31 | 0 |
| rs706785 | eQTLGen | eQTLGen_cis_eQTLs | *LIMA1* | C | 10.77 | 4.6E-27 | 0 |
| rs706785 | eQTLcatalogue | TwinsUK_ge_fat | *RACGAP1* | C | -0.45 | 3.7E-24 | 1.9E-20 |
| rs706785 | eQTLcatalogue | Fairfax_2014_IFN24 | *RACGAP1* | C | -0.19 | 3.6E-18 | 1.8E-14 |
| rs706785 | GTEx/v8 | Adipose_Subcutaneous | *RACGAP1* | A | -0.36 | 3.6E-15 | 8.1E-17 |
| rs706785 | eQTLGen | eQTLGen_cis_eQTLs | *COX14* | C | 7.50 | 6.3E-14 | 0 |
| rs706785 | eQTLcatalogue | Fairfax_2014_IFN24 | *LIMA1* | C | -0.28 | 5.0E-11 | 2.5E-07 |
| rs706785 | GTEx/v7 | Adipose_Subcutaneous | *RACGAP1* | A | -0.40 | 1.3E-10 | 4.8E-11 |
| rs706785 | BIOSQTL | BIOS_eQTL_geneLevel | *LIMA1* | C | 6.29 | 3.2E-10 | 0 |
| rs706785 | eQTLcatalogue | Fairfax_2014_naive | *RACGAP1* | C | -0.13 | 4.4E-10 | 2.2E-06 |
| rs706785 | BIOSQTL | BIOS_eQTL_geneLevel | *COX14* | C | 5.97 | 2.4E-09 | 6.1E-06 |
| rs706785 | GTEx/v7 | Breast_Mammary_Tissue | *RACGAP1* | A | -0.48 | 1.2E-07 | 5.6E-07 |
| rs706785 | GTEx/v8 | Brain_Nucleus_accumbens_basal_ganglia | *RP4-605O3.4* | A | 0.58 | 1.5E-07 | 2.1E-14 |
| rs706785 | eQTLGen | eQTLGen_cis_eQTLs | *SMARCD1* | C | 5.17 | 2.3E-07 | 7.7E-04 |
| rs706785 | GTEx/v6 | Adipose_Subcutaneous | *RACGAP1* | A | -5.27 | 2.8E-07 | 2.7E-04 |
| rs706785 | GTEx/v8 | Breast_Mammary_Tissue | *RACGAP1* | A | -0.30 | 6.2E-07 | 5.4E-06 |
| rs706785 | GTEx/v8 | Muscle_Skeletal | *RP4-605O3.4* | A | 0.32 | 9.6E-07 | 1.8E-20 |
| rs706785 | BIOSQTL | BIOS_eQTL_geneLevel | *RACGAP1* | C | 4.80 | 1.6E-06 | 1.3E-03 |
| rs706785 | GTEx/v7 | Whole_Blood | *LIMA1* | A | -0.23 | 6.1E-06 | 4.3E-05 |
| rs706785 | eQTLcatalogue | Fairfax_2014_naive | *LIMA1* | C | -0.16 | 8.5E-06 | 4.2E-02 |
| rs706785 | GTEx/v8 | Adipose_Visceral_Omentum | *RACGAP1* | A | -0.25 | 1.2E-05 | 3.2E-06 |
| rs706785 | GTEx/v7 | Muscle_Skeletal | *RP4-605O3.4* | A | 0.36 | 1.5E-05 | 3.0E-08 |
| rs706785 | GTEx/v8 | Breast_Mammary_Tissue | *AQP6* | A | -0.31 | 1.6E-05 | 1.0E-04 |
| rs706785 | GTEx/v6 | Cells_Transformed_fibroblasts | *FMNL3* | A | -4.31 | 2.4E-05 | 2.5E-02 |
| rs706785 | GTEx/v8 | Esophagus_Gastroesophageal_Junction | *RP4-605O3.4* | A | 0.43 | 2.5E-05 | 2.6E-16 |
| rs706785 | GTEx/v7 | Heart_Left_Ventricle | *RP4-605O3.4* | A | 0.41 | 3.3E-05 | 5.2E-09 |
| rs706785 | GTEx/v8 | Adipose_Visceral_Omentum | *AQP6* | A | -0.31 | 3.3E-05 | 1.3E-02 |
| rs706785 | GTEx/v8 | Heart_Left_Ventricle | *RP4-605O3.4* | A | 0.31 | 7.1E-05 | 4.6E-18 |
| rs706785 | GTEx/v8 | Nerve_Tibial | *CERS5* | A | 0.24 | 7.8E-05 | 2.7E-31 |
| rs706785 | GTEx/v8 | Thyroid | *FAM186A* | A | 0.23 | 1.0E-04 | 1.8E-03 |
| rs706785 | GTEx/v8 | Pancreas | *COX14* | A | -0.25 | 1.3E-04 | 2.6E-06 |
| rs706785 | GTEx/v8 | Cells_Cultured_fibroblasts | *ASIC1* | A | 0.15 | 1.4E-04 | 1.0E-15 |
| rs706785 | EyeGEx | EyeGEx | *SMARCD1* | C | 0.51 | 1.4E-04 | 4.9E-02 |
| rs706785 | GTEx/v8 | Cells_Cultured_fibroblasts | *COX14* | A | -0.16 | 1.5E-04 | 1.9E-12 |
| rs694851 | eQTLGen | eQTLGen_cis_eQTLs | *CERS5* | C | -16.06 | 4.9E-58 | 0 |
| rs694851 | eQTLGen | eQTLGen_cis_eQTLs | *LIMA1* | C | 11.36 | 6.6E-30 | 0 |
| rs694851 | eQTLGen | eQTLGen_cis_eQTLs | *RACGAP1* | C | 11.07 | 1.7E-28 | 0 |
| rs694851 | eQTLcatalogue | TwinsUK_ge_fat | *RACGAP1* | C | -0.45 | 3.7E-24 | 1.9E-20 |
| rs694851 | eQTLcatalogue | Fairfax_2014_IFN24 | *RACGAP1* | C | -0.19 | 4.9E-18 | 2.5E-14 |
| rs694851 | eQTLGen | eQTLGen_cis_eQTLs | *COX14* | C | 8.00 | 1.2E-15 | 0 |
| rs694851 | GTEx/v8 | Adipose_Subcutaneous | *RACGAP1* | G | -0.36 | 3.9E-15 | 8.1E-17 |
| rs694851 | eQTLcatalogue | Fairfax_2014_IFN24 | *LIMA1* | C | -0.28 | 3.2E-11 | 1.6E-07 |
| rs694851 | GTEx/v7 | Adipose_Subcutaneous | *RACGAP1* | G | -0.40 | 1.3E-10 | 4.8E-11 |
| rs694851 | eQTLcatalogue | Fairfax_2014_naive | *RACGAP1* | C | -0.13 | 1.7E-10 | 8.6E-07 |
| rs694851 | BIOSQTL | BIOS_eQTL_geneLevel | *LIMA1* | C | 6.36 | 2.0E-10 | 0 |
| rs694851 | BIOSQTL | BIOS_eQTL_geneLevel | *COX14* | C | 5.95 | 2.7E-09 | 6.0E-06 |
| rs694851 | GTEx/v7 | Breast_Mammary_Tissue | *RACGAP1* | G | -0.48 | 1.2E-07 | 5.6E-07 |
| rs694851 | GTEx/v8 | Brain_Nucleus_accumbens_basal_ganglia | *RP4-605O3.4* | G | 0.58 | 1.5E-07 | 2.1E-14 |
| rs694851 | GTEx/v6 | Adipose_Subcutaneous | *RACGAP1* | G | -5.30 | 2.4E-07 | 2.7E-04 |
| rs694851 | GTEx/v8 | Breast_Mammary_Tissue | *RACGAP1* | G | -0.30 | 6.2E-07 | 5.4E-06 |
| rs694851 | GTEx/v8 | Muscle_Skeletal | *RP4-605O3.4* | G | 0.31 | 1.1E-06 | 1.8E-20 |
| rs694851 | BIOSQTL | BIOS_eQTL_geneLevel | *RACGAP1* | C | 4.87 | 1.1E-06 | 9.5E-04 |
| rs694851 | eQTLGen | eQTLGen_cis_eQTLs | *SMARCD1* | C | 4.78 | 1.8E-06 | 5.3E-03 |
| rs694851 | GTEx/v7 | Whole_Blood | *LIMA1* | G | -0.23 | 5.0E-06 | 4.3E-05 |
| rs694851 | eQTLcatalogue | Fairfax_2014_naive | *LIMA1* | C | -0.16 | 6.1E-06 | 3.0E-02 |
| rs694851 | eQTLcatalogue | Fairfax_2012_B-cell_CD19 | *CERS5* | C | 0.16 | 6.6E-06 | 3.3E-02 |
| rs694851 | GTEx/v8 | Adipose_Visceral_Omentum | *RACGAP1* | G | -0.26 | 8.0E-06 | 3.2E-06 |
| rs694851 | GTEx/v8 | Breast_Mammary_Tissue | *AQP6* | G | -0.31 | 1.6E-05 | 1.0E-04 |
| rs694851 | GTEx/v7 | Muscle_Skeletal | *RP4-605O3.4* | G | 0.35 | 1.9E-05 | 3.0E-08 |
| rs694851 | GTEx/v8 | Adipose_Visceral_Omentum | *AQP6* | G | -0.32 | 2.4E-05 | 1.3E-02 |
| rs694851 | GTEx/v8 | Esophagus_Gastroesophageal_Junction | *RP4-605O3.4* | G | 0.42 | 3.1E-05 | 2.6E-16 |
| rs694851 | GTEx/v7 | Heart_Left_Ventricle | *RP4-605O3.4* | G | 0.41 | 3.4E-05 | 5.2E-09 |
| rs694851 | GTEx/v8 | Heart_Left_Ventricle | *RP4-605O3.4* | G | 0.31 | 7.1E-05 | 4.6E-18 |
| rs694851 | GTEx/v8 | Thyroid | *FAM186A* | G | 0.23 | 7.3E-05 | 1.8E-03 |
| rs694851 | GTEx/v8 | Nerve_Tibial | *CERS5* | G | 0.24 | 7.8E-05 | 2.7E-31 |
| rs694851 | GTEx/v8 | Cells_Cultured_fibroblasts | *ASIC1* | G | 0.15 | 1.4E-04 | 1.0E-15 |
| rs694851 | EyeGEx | EyeGEx | *SMARCD1* | C | 0.51 | 1.4E-04 | 4.8E-02 |
| rs694851 | GTEx/v8 | Cells_Cultured_fibroblasts | *COX14* | G | -0.16 | 1.5E-04 | 1.9E-12 |
| rs694851 | GTEx/v8 | Adipose_Subcutaneous | *AQP6* | G | -0.30 | 2.1E-04 | 2.4E-02 |
| rs615382 | eQTLGen | eQTLGen_cis_eQTLs | *CERS5* | G | -15.77 | 5.2E-56 | 0 |
| rs615382 | eQTLGen | eQTLGen_cis_eQTLs | *RACGAP1* | G | 11.54 | 8.2E-31 | 0 |
| rs615382 | eQTLGen | eQTLGen_cis_eQTLs | *LIMA1* | G | 10.72 | 8.5E-27 | 0 |
| rs615382 | eQTLcatalogue | TwinsUK_ge_fat | *RACGAP1* | G | -0.45 | 3.7E-24 | 1.9E-20 |
| rs615382 | eQTLcatalogue | Fairfax_2014_IFN24 | *RACGAP1* | G | -0.19 | 5.9E-18 | 2.9E-14 |
| rs615382 | eQTLGen | eQTLGen_cis_eQTLs | *COX14* | G | 8.29 | 1.2E-16 | 0 |
| rs615382 | GTEx/v8 | Adipose_Subcutaneous | *RACGAP1* | A | -0.37 | 2.8E-15 | 8.1E-17 |
| rs615382 | eQTLcatalogue | Fairfax_2014_IFN24 | *LIMA1* | G | -0.28 | 2.8E-11 | 1.4E-07 |
| rs615382 | GTEx/v7 | Adipose_Subcutaneous | *RACGAP1* | A | -0.40 | 1.7E-10 | 4.8E-11 |
| rs615382 | BIOSQTL | BIOS_eQTL_geneLevel | *LIMA1* | G | 6.38 | 1.8E-10 | 0 |
| rs615382 | eQTLcatalogue | Fairfax_2014_naive | *RACGAP1* | G | -0.13 | 2.2E-10 | 1.1E-06 |
| rs615382 | BIOSQTL | BIOS_eQTL_geneLevel | *COX14* | G | 5.92 | 3.2E-09 | 6.0E-06 |
| rs615382 | GTEx/v7 | Breast_Mammary_Tissue | *RACGAP1* | A | -0.52 | 1.5E-08 | 5.6E-07 |
| rs615382 | GTEx/v8 | Brain_Nucleus_accumbens_basal_ganglia | *RP4-605O3.4* | A | 0.58 | 1.5E-07 | 2.1E-14 |
| rs615382 | GTEx/v6 | Adipose_Subcutaneous | *RACGAP1* | A | -5.32 | 2.2E-07 | 2.7E-04 |
| rs615382 | GTEx/v8 | Muscle_Skeletal | *RP4-605O3.4* | A | 0.32 | 9.2E-07 | 1.8E-20 |
| rs615382 | GTEx/v8 | Breast_Mammary_Tissue | *RACGAP1* | A | -0.30 | 9.5E-07 | 5.4E-06 |
| rs615382 | BIOSQTL | BIOS_eQTL_geneLevel | *RACGAP1* | G | 4.87 | 1.1E-06 | 9.5E-04 |
| rs615382 | eQTLGen | eQTLGen_cis_eQTLs | *SMARCD1* | G | 4.88 | 1.1E-06 | 3.3E-03 |
| rs615382 | GTEx/v7 | Whole_Blood | *LIMA1* | A | -0.23 | 4.5E-06 | 4.3E-05 |
| rs615382 | eQTLcatalogue | Fairfax_2014_naive | *LIMA1* | G | -0.16 | 5.9E-06 | 3.0E-02 |
| rs615382 | eQTLcatalogue | Fairfax_2012_B-cell_CD19 | *CERS5* | G | 0.16 | 6.1E-06 | 3.1E-02 |
| rs615382 | GTEx/v7 | Muscle_Skeletal | *RP4-605O3.4* | A | 0.36 | 1.5E-05 | 3.0E-08 |
| rs615382 | GTEx/v8 | Adipose_Visceral_Omentum | *AQP6* | A | -0.33 | 1.7E-05 | 1.3E-02 |
| rs615382 | GTEx/v8 | Adipose_Visceral_Omentum | *RACGAP1* | A | -0.25 | 2.1E-05 | 3.2E-06 |
| rs615382 | GTEx/v8 | Esophagus_Gastroesophageal_Junction | *RP4-605O3.4* | A | 0.43 | 2.5E-05 | 2.6E-16 |
| rs615382 | GTEx/v6 | Cells_Transformed_fibroblasts | *FMNL3* | A | -4.28 | 2.8E-05 | 2.5E-02 |
| rs615382 | GTEx/v7 | Heart_Left_Ventricle | *RP4-605O3.4* | A | 0.41 | 3.3E-05 | 5.2E-09 |
| rs615382 | GTEx/v8 | Breast_Mammary_Tissue | *AQP6* | A | -0.30 | 4.1E-05 | 1.0E-04 |
| rs615382 | GTEx/v8 | Heart_Left_Ventricle | *RP4-605O3.4* | A | 0.31 | 7.1E-05 | 4.6E-18 |
| rs615382 | GTEx/v8 | Nerve_Tibial | *CERS5* | A | 0.24 | 7.6E-05 | 2.7E-31 |
| rs615382 | GTEx/v8 | Cells_Cultured_fibroblasts | *ASIC1* | A | 0.15 | 1.1E-04 | 1.0E-15 |
| rs615382 | GTEx/v8 | Thyroid | *FAM186A* | A | 0.23 | 1.1E-04 | 1.8E-03 |
| rs615382 | EyeGEx | EyeGEx | *SMARCD1* | G | 0.52 | 1.3E-04 | 4.5E-02 |
| rs615382 | GTEx/v8 | Pancreas | *COX14* | A | -0.25 | 1.3E-04 | 2.6E-06 |
| rs615382 | GTEx/v8 | Cells_Cultured_fibroblasts | *COX14* | A | -0.17 | 1.4E-04 | 1.9E-12 |

***** The information of eQTL summary statistics were retrieved from several eQTL studies and databases using FUMA (*Nat Commun* 2017;8:1826; https://fuma.ctglab.nl/tutorial#eQTLs).

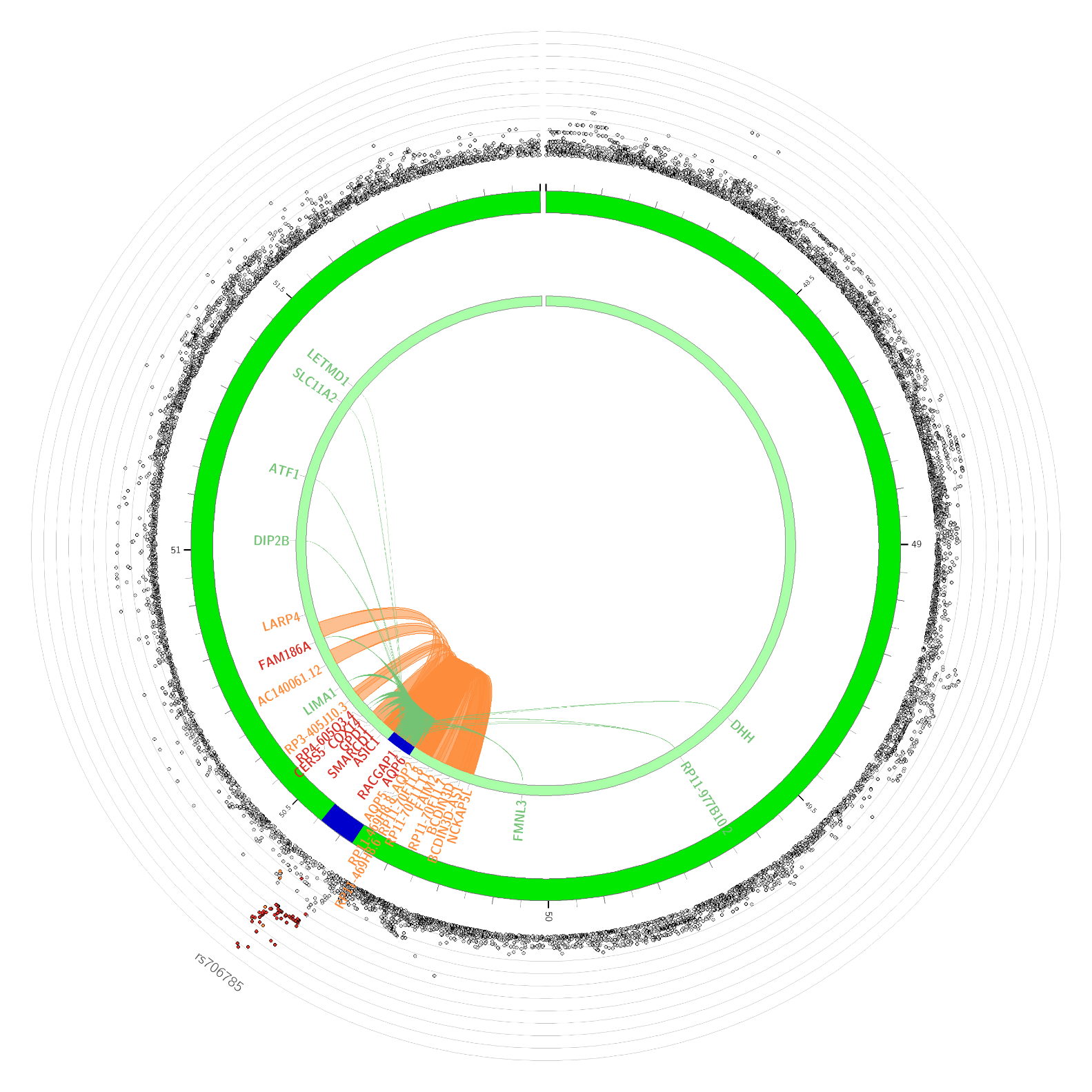

**Supplementary Figure 1.** The Circos plot for the *RACGAP1* locus displaying the GWAS locus (blue), its chromatin interaction (orange loops), and reported genes (green loops) across tissues. The outer layer in the plot represents the GWAS *P* values in log scale. The chromatin interaction was retrieved from previous studies and databases using FUMA (*Nat Commun* 2017;8:1826; https://fuma.ctglab.nl/tutorial#chromatin-interactions).
